# Effect of Eligibility Criteria on Patients’ Survival and Serious Adverse Events in Colorectal Cancer Drug Trials

**DOI:** 10.1101/2024.04.03.24305265

**Authors:** Chang Wang, Aokun Chen, Xing He, Patrick Balian, Thomas J. George, Fei Wang, Jiang Bian, Yi Guo

## Abstract

This study investigates the impact of clinical trial eligibility criteria on patient survival and serious adverse events (SAEs) in colorectal cancer (CRC) drug trials using real-world data. We utilized the OneFlorida+ network’s data repository, conducting a retrospective analysis of CRC patients receiving FDA-approved first-line metastatic treatments. Propensity score matching created balanced case-control groups, which were evaluated using survival analysis and machine learning algorithms to assess the effects of eligibility criteria. Our study included 68,375 patients, with matched case-control groups comprising 1,126 patients each. Survival analysis revealed ethnicity and race, along with specific medical history (eligibility criteria), as significant survival outcome predictors. Machine learning models, particularly the XgBoost regressor, were employed to analyze SAEs, indicating that age and study groups were notable factors in SAEs occurrence. The study’s findings highlight the importance of considering patient demographics and medical history in CRC trial designs.

## Introduction

Colorectal cancer (CRC) is a serious global health concern, standing as the third most prevalent cancer and the second leading cause of cancer-related deaths worldwide.^1^ In 2020, the global burden of CRC was marked by an estimated 1.9 million new cases and over 930,000 deaths.^1^ By 2040, projections indicate a significant rise in both incidence and mortality rates: new cases are expected to increase by 63%, reaching 3.2 million annually, and deaths are anticipated to surge by 73%, amounting to 1.6 million.^1^ In the United States, CRC ranks as the third and fourth most common cause of cancer-related fatalities among males and females, respectively.^2^ Combining the death tolls from both genders, it becomes the second most common cause of cancer-related deaths. The American Cancer Society projects approximately 106,590 new cases of colon cancer and 46,220 new cases of rectal cancer in the U.S. in 2024.^2^ These staggering statistics underscore an urgent need for effective CRC treatments, predominantly evaluated through clinical trials. Randomized controlled trials (RCTs) are the gold standard for assessing the safety and efficacy of new treatments.^3^ However, the eligibility criteria for these trials, crucial for ensuring patient safety and data integrity, can significantly influence their outcomes. Restrictive criteria may hinder participant recruitment and limit the findings’ applicability to the broader CRC patient population, thereby limiting the external validity of clinical trial outcomes.^4,5^

Against this backdrop, real-world data (RWD)—encompassing health information regularly gathered from diverse sources beyond conventional research settings—emerges as a pivotal resource. RWD transcends the limitations of traditional epidemiological research and clinical trials, offering a vast, cost-effective data source for healthcare insights.^6,7^ This study leverages RWD from electronic health records (EHRs) to explore the impact of eligibility criteria on patient survival and serious adverse events (SAEs) in CRC drug trials. Utilizing the OneFlorida+ network, part of the national Patient-Centered Outcomes Research Network (PCORnet), we access a comprehensive RWD repository of over 21.19 million patients from Florida, Georgia, and Alabama.^8^ This rich dataset, derived from EHRs, insurance claims, and cancer registries, provides a unique platform for assessing the real-world implications of clinical trial eligibility criteria on patient outcomes.

Our research employs the OneFlorida+ database for a retrospective analysis of CRC patients receiving first-line metastatic treatments approved by the US Food and Drug Administration (FDA). By replicating the conditions of phase III clinical trials within this real-world cohort, we aim to illuminate how eligibility criteria affect patient survival and SAEs. The incorporation of advanced machine learning (ML) techniques allows for an in-depth examination of the relationship between trial-specific criteria and patient outcomes, uncovering potential disparities between clinical research and actual clinical practice. By bridging the gap between the rigor of clinical trials and the realities of clinical practice, our study aspires to refine future trial designs and regulatory policies. The goal is to ensure that advancements in CRC treatment are effectively translated into improved outcomes for a diverse patient population, addressing the critical global burden of colorectal cancer.

## Methods

### Data source

The OneFlorida+ network, a large CRN in the national PCORnet supported by the Patient-Centered Outcomes Research Institute, provided the real-world EHR data used in this study. The OneFlorida+ data trust is a Health Insurance Portability and Accountability Act (HIPAA)-restricted data collection that includes comprehensive patient demographic and clinical information, such as demographics, encounters, diagnoses, procedures, vitals, prescriptions, and laboratory findings. The Institutional Review Board (IRB) of the University of Florida approved this study.

We conducted a comprehensive analysis of four phase III clinical trials involving the first-line treatment for metastatic CRC, as approved by the US FDA. Patients with CRC were identified using the International Classification of Diseases (ICD) codes (153, 154, 159.0, C18, C19, C20, C21, and C26.0). Patients with incomplete demographic information were excluded. This process yielded an initial cohort of 68,375 patients. The index date was defined as the day on which a patient received the first CRC diagnosis. We used the National Drug Code (NDC), RxNorm Concept Unique Identifier (RxCUI), and Healthcare Common Procedure Coding System (HCPCS) codes to ascertain the utilization of CRC-specific medications.

### Case and Control Definitions

Given that each of the four trials we selected investigated a combination therapy incorporating the FOLFIRI regimen (5-FU, leucovorin or levoleucovorin, and irinotecan) with a unique single agent as an intervention, we defined our initial case-cohort to include patients undergoing such combination treatments. Specifically, the therapies under analysis involved FOLFIRI in conjunction with one of the following agents: panitumumab (NCT00339183), cetuximab (NCT00154102), sunitinib (NCT00339183), or ramucirumab (NCT01183780). Considering the uniform application of FOLFIRI alone as the control in all selected trials, our initial control group comprised patients treated exclusively with the FOLFIRI regimen, without the addition of any other agents. Drug administration had to occur post-index date, and we included various formulations such as injectables, tablets, and capsules. To emulate the conditions of CRC clinical trials using EHR data, propensity score matching (PSM) was employed to align control patients with case patients based on age, sex, race, and ethnicity, ensuring a 1:1 ratio. The index dates for matched controls were mandated to be within a one-year range of the case index date. Following the matching process, we obtained 1,223 case-control pairs. Then, we eliminated the outliers of SAEs via Z-score analysis; the cohorts were finalized at 1,126 patients each. The patient selection flowchart is shown in **Figure 1**.

**Figure 1.**
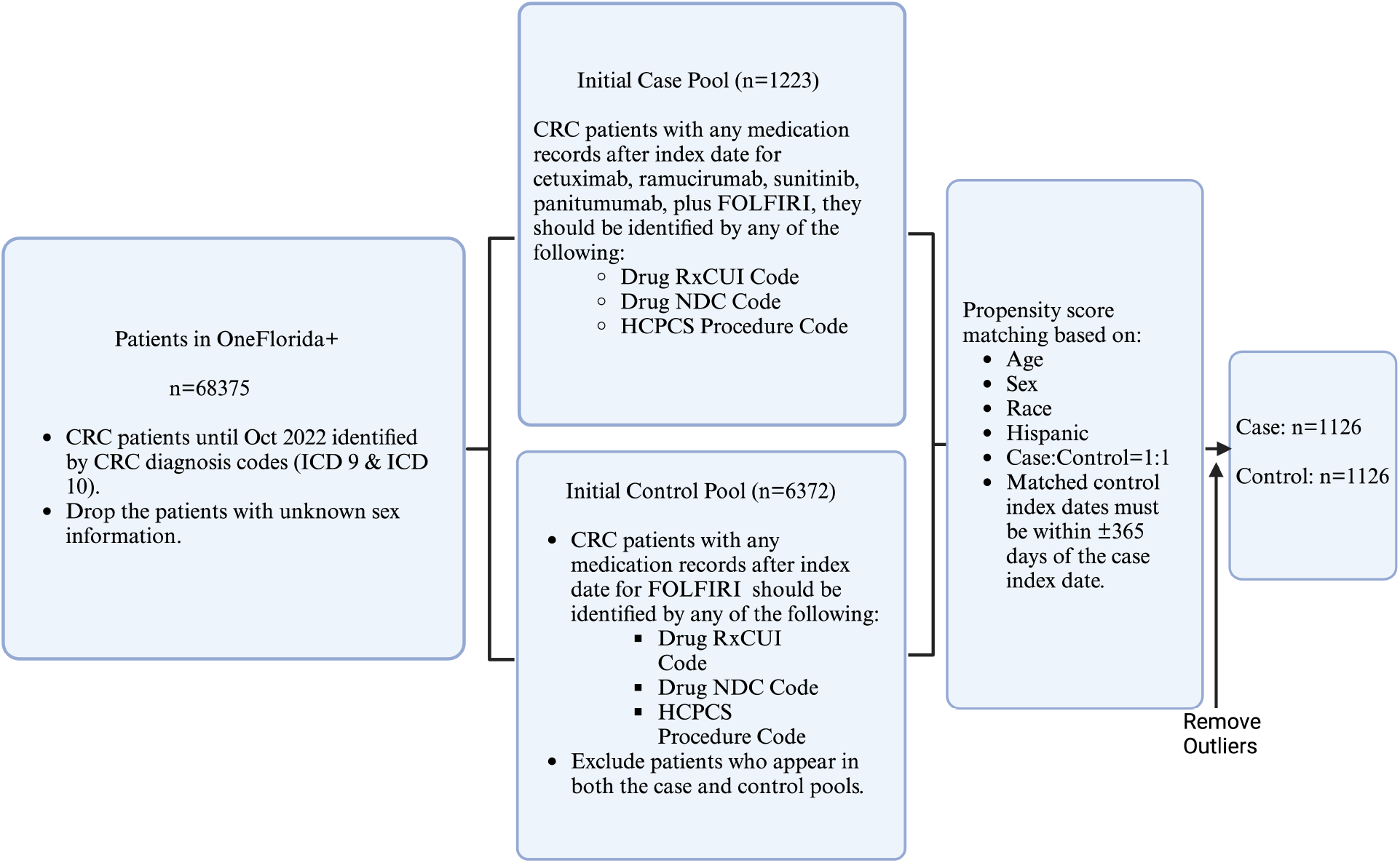
Overall Study design and selection of the population.

### Study Outcomes

Our study analyzed two outcomes of interest: the overall survival of the emulated CRC cohort and the SAEs. The overall survival of the cancer patients was observed over a maximum five-year follow-up period, starting from the first incidence of CRC to the death of any cause. If the event of interest (in this case, death of any cause) had not occurred by the end of the observation period, a censor event was assigned. Regarding the SAE analysis, the SAE was evaluated based on the SAE events experienced by the emulated study cohort. We measured the incidence of the therapy-related SAEs reported by the 4 clinical trials inside our SAE observation window. As shown in **Figure 2**, the observation window for SAEs spanned across the date of the initial drug administration and 30 days after the date of the final drug administration, identified by the last instance of drug dispensation or administration. We used the International Classification of Diseases (ICD-9/10-CM) codes to identify the SAEs from the OneFlorida+ EHR and extract the count of unique SAE-related encounters.

**Figure 2.**
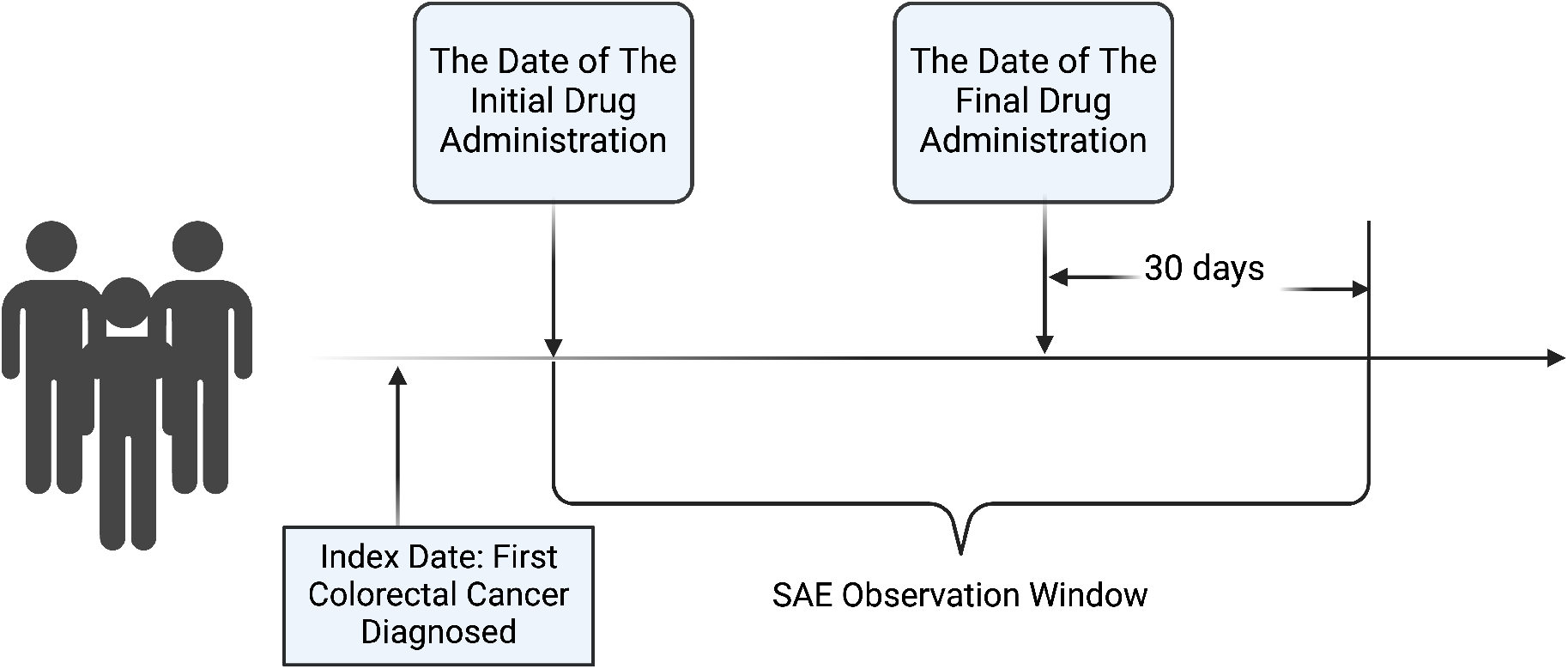
SAE observation window.

### Survival Analysis

Our analysis assessed survival outcomes using both non-parametric and Cox proportional hazards methodologies. Initially, Kaplan-Meier (KM) survival curves were constructed to facilitate direct comparison between the two emulated study groups, with the log-rank test determining statistical differences in their survival distributions.

Subsequently, we developed a Cox proportional hazards model further to investigate the influence of specific study traits as predictors of survival, concurrently adjusting for demographic factors including age, sex, race, and ethnicity. Before constructing the Cox model, we removed the highly correlated variables using the variance inflation factor (VIF). In this process, the variables with the event more frequently observed among the selected CRC patients were retained. For instance, the variables “history of another primary cancer within 5 years before randomization” and “history of another primary cancer within 3 years before randomization” were closely related. We kept the former as the event ‘history of another primary cancer within 5 years’ was more frequently observed in our cohort. Furthermore, we enhanced the model’s predictive power and generalizability by excluding the infrequently observed study traits. The complete list of removed and retained eligibility criteria can be found in **Supplement Table 1**.

### SAE analysis with Machine Learning Algorithms

We combined ML methods with SHapley Additive exPlanations (SHAP)^9^ to explore the relationship between the SAE and the study traits adjusted for demographic factors. As the first step, we identified the best-performing machine learning model from a series of models widely adopted in medical research: support vector regressor (SVR), extreme gradient boost (XgBoost) regressor, and Adaboost regressor (Adaboost). SVR is a supervised machine learning technique based on the Vapnik-Chervonenkis (support vector) theory.^10^ Fitted with a learned margin, it could potentially be more accurate and generalizable compared to other regressors. The XgBoost regressor is a gradient boost-based ensemble learning technique that, for high-efficiency tree split selection, substitutes an effective approximation approach for the exact greedy algorithm.^11^ In clinical research, the XgBoost regressor is known for being more effective and accurate than other tree-based machine-learning techniques.^12^ Adaboost is an ensemble learning technique that enhances performance over difficult-to-predict subgroups by using relative error and weighting techniques.^13^ To fit the ML models, we randomly divided the emulated study groups into training and testing sets at a 4:1 ratio. The training set was used to train the three regressors, while the test set was used for evaluation. Based on its superior performance in evaluations, we chose the XgBoost regressor for further study.

With the best algorithm identified, we computed SHAP values to assess the impact of each study trait on SAE within our emulated CRC study groups. **Equation (1)** was used to determine each SHAP value based on the published XgBoost regressor models. In essence, the SHAP value describes each predictor’s contribution to the change in model output as follows:

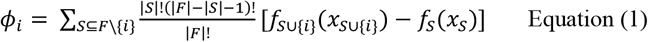

Where i is a single predictor (i.e., the study trait in this study), F is the set of all predictors, S is a subset of F, and *x*_*S*_ is the predictors in S.

## Results

### CRC Eligibility Criteria and Study Trait Pool

The four CRC trials contained a total of 60 eligibility criteria. However, not all conditions described by eligibility criteria could be extracted from structured EHR, i.e., computable. Also, one eligibility criterion could contain multiple conditions, requiring further decomposition into study traits on a single condition for computing and representation. As an example, the eligibility criterion “History of the presence of brain metastasis, spinal cord compression carcinomatous meningitis or leptomeningeal disease” was decomposed into 2 study traits with brain metastasis, spinal cord compression carcinomatous meningitis or leptomeningeal disease. Among the 60 eligibility criteria, we identified 20 computable criteria and 40 incomputable criteria. The trials NCT01183780 and NCT00339183 contained the most computable eligibility criteria, with 8 computable eligibility criteria, each that could be further divided into 13 and 9 study traits, respectively. Both NCT00457691 and NCT00154102 trials had a minimum of 2 computable eligibility criteria. We combined the study traits from four CRC trials and formed a study trait pool of 22 distinct, computable study traits as the study predictors. These predictors were coded as binary variables representing whether a patient had the condition indicated by a particular study trait (0 = condition not present or 1 = condition present). **Supplement Table 1** presents the computable eligibility criteria alongside their corresponding decomposed study traits for each trial.

### Demographics of CRC Patients

We extracted a cohort of CRC patients from OneFlorida+ between 2012 and 2022. The demographic information for the 68,375 CRC patients is summarized in **Table 1**. The average age of our CRC cohort was 63.74 years, with a standard deviation of 14.1 years. An almost equal gender distribution was observed (female 50.2%). Regarding ethnicity, 21.8% of patients were Hispanic, 68.5% were non-Hispanic, and the remaining 9.8% were Other or Unknown. The CRC cohort included 63.2% White patients, 16.2% Black patients, and 20.5% patients with other or unknown races.

**Table 1.** Characteristics of total CRC patients, drug used case group, and matched control group.

| Variables | Total CRC patients<br>n=68,375 | Case group<br>n= 1126 | Control group<br>n=1126 | P-value for<br>study groups |
| --- | --- | --- | --- | --- |
| Age, Mean (SD) | 63.74 (14.1) | 56.22 (11.1) | 56.16 (10.8) | 0.84 |
| Sex |  |  |  | 1.00 |
| Female | 34,295 (50.2 %) | 476 (42.3 %) | 476 (42.3%) |  |
| Male | 34,080 (49.8 %) | 650 (57.7%) | 650 (57.7%) |  |
| Hispanic |  |  |  | 0.56 |
| Yes | 14,886 (21.8 %) | 241 (21.4 %) | 226 (20.1 %) |  |
| No | 46,805 (68.5 %) | 743 (66.0%) | 767 (68.1 %) |  |
| Other or Unknown | 6684 (9.8%) | 142 (12.6 %) | 133 (11.8%) |  |
| Race |  |  |  | 0.66 |
| White | 43,227 (63.2 %) | 637 (56.6 %) | 643 (57.1 %) |  |
| Black | 11,101 (16.2 %) | 200 (17.8 %) | 211 (18.7%) |  |
| Other or Unknown | 14,047 (20.5%) | 289 (25.7 %) | 272 (24.2 %) |  |

Following PSM, we established the matched case and control groups, each comprising 1,126 patients, as detailed in **Table 1**. The mean age was 56.22 years (SD = 11.1) in the case group and 56.16 years (SD = 10.8) in the control group. The sex distribution was identical in both groups (p = 1.00), with females representing 42.3% (n = 476) and males 57.7% (n = 650). Ethnicity comparison showed a slight difference (p = 0.56); in the case group, 21.4% (n = 241) were Hispanic, 66.0% (n = 743) non-Hispanic, and 12.61% (n = 142) were Other or Unknown. In contrast, the control group had 20.1% (n = 226) Hispanic, 68.1% (n = 767) non-Hispanic, and 11.8% (n = 133) Other or Unknown, indicating a closely matched ethnic composition. The racial composition also showed a close match between the two groups. In the case group, Whites constituted 56.6% (n = 637), Blacks 17.8% (n = 200), and Other or Unknown 25.7% (n = 289). The control group had 57.1% (n = 643) Whites, 18.7% (n = 211) Blacks, and 24.2% (n = 272) Other or Unknown (p = 0.66). The p-values for age, sex distribution, ethnicity, and racial composition comparisons between the case and control groups were all above the conventional significance threshold of 0.05, indicating no statistically significant differences and suggesting well-balanced study groups.

### Survival Analysis

**Figure 3** presents the Kaplan-Meier survival curves for two patient cohorts monitored over a 5-year period, stratified into case (blue) and control (orange). The survival probability is plotted on the y-axis, while the x-axis denotes the time to event or censor in years. Initially, both groups demonstrated nearly identical survival probabilities. As time progresses, a slight divergence emerges between the curves, with the case group displaying a marginally higher survival probability compared to the control group. Despite this observed divergence, the log-rank test did not reveal a statistically significant difference in survival probabilities between the case and control groups (p = 0.147).

**Figure 3.**
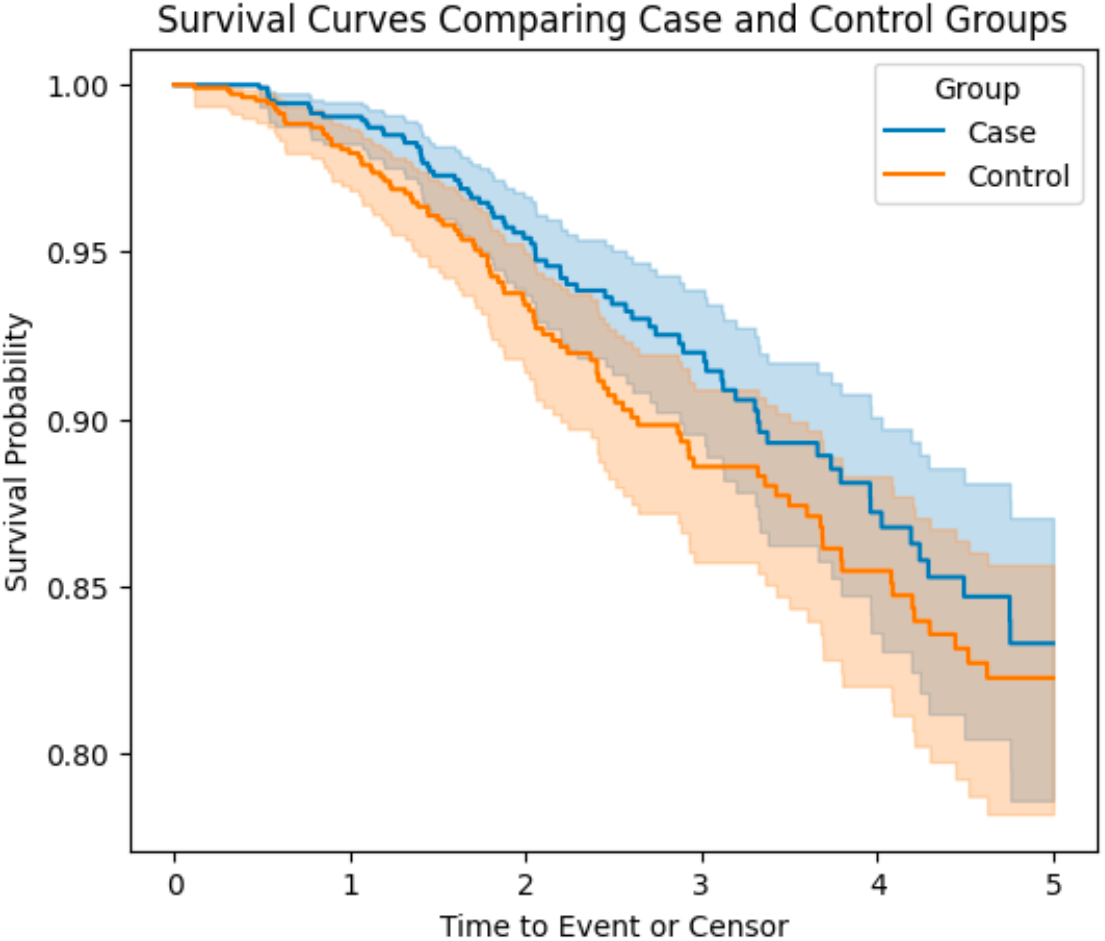
Kaplan-Meier survival curves for emulated case and control cohorts.

However, subsequent examination through a Cox proportional hazards model identified four predictors with statistically significant impacts on the time to death (**Table 2**). Ethnicity emerged as a significant determinant, with individuals identifying as Hispanic demonstrating a 54% increase in the hazard of death compared to non-Hispanic individuals (Hazard Ratio [HR] = 1.54; 95% Confidence Interval [CI]: 1.01-2.35; p = 0.04). Similarly, the racial category of ‘Other or Unknown,’ when compared to Black individuals, was associated with a substantial 70% decrease in the hazard of death (HR = 0.30; 95% CI: 0.14-0.66; p < 0.005). A notable increase in the hazard of death was observed among patients with a history of another primary cancer within 5 years preceding randomization, with an HR indicating more than a twofold increase (HR = 2.06; 95% CI: 1.51-2.83; p<0.005). Additionally, the occurrence of myocardial infarction within 12 months prior to randomization was linked to a markedly elevated hazard of death, nearly sixfold higher compared to those without such a history (HR = 5.92; 95% CI: 2.40-14.61; p < 0.005).

**Table 2.** Cox proportional hazard model results.

| Variables | Hazard Ratio (95% CI) | p-value |
| --- | --- | --- |
| Age | 1.00 (0.98, 1.01) | 0.69 |
| SEX: Male vs. Female | 1.03 (0.76, 1.41) | 0.84 |
| HISPANIC: Other or Unknown vs. No | 0.28 (0.06, 1.34) | 0.11 |
| Hispanic: Yes vs. No | 1.54 (1.01, 2.35) | 0.04 |
| Race: Other or Unknown vs. Black | 0.30 (0.14, 0.66) | <0.005 |
| Race: White vs. Black | 0.80 (0.55, 1.16) | 0.24 |
| Group: Case vs. Control | 0.78 (0.57, 1.06) | 0.11 |
| History of another primary cancer within 5 years of randomization | 2.06 (1.51, 2.82) | <0.005 |
| Inflammatory bowel disease | 1.73 (0.76, 3.96) | 0.19 |
| HIV | 0.59 (0.18, 1.99) | 0.40 |
| HBV or HCV | 1.28 (0.46, 3.59) | 0.64 |
| History of interstitial lung disease | 0.94 (0.13, 6.98) | 0.95 |
| Prior irinotecan therapy | 1.64 (0.51, 5.25) | 0.41 |
| Receipt of bevacizumab $\leq 28$ days prior to randomization | 0.72 (0.29, 1.79) | 0.48 |
| Experience of myocardial infarction $\leq 12$ months prior to randomization | 5.92 (2.40, 14.61) | <0.005 |
| Experience of transient ischemic attack $\leq 12$ months prior to randomization | 1.61 (0.35, 7.51) | 0.54 |
| Experience of cerebrovascular accident $\leq 12$ months prior to randomization | 2.93 (0.63, 13.65) | 0.17 |
| Bowel obstruction occur at randomization day | 0.83 (0.52, 1.33) | 0.44 |
| History of diarrhea | 0.98 (0.57, 1.68) | 0.93 |
| Cirrhosis (any degree) | 1.82 (0.60, 5.50) | 0.29 |

These findings underscore the significance of ethnic and racial background, as well as specific medical histories, in influencing survival outcomes among the study population. The differential impacts highlighted by these predictors warrant further investigation to understand their underlying mechanisms and potential implications for patient management and care strategies.

### Machine Learning Algorithms and SHAP Value

Our SHAP value analysis presents a comprehensive visualization of the feature impact on the XgBoost model’s predictions concerning SAEs in metastatic CRC patients. The results elucidate the multifaceted nature of clinical trial exclusion criteria and their influence on SAE occurrence.

**Figure 4** presents the SHAP value summary plot, illustrating the impact of each study trait on SAE. The SHAP summary plot illustrates that age and study groups exerted the greatest impact on the model’s predictions, with higher SHAP values indicating a strong association with increased risk of SAEs. Notably, patient age showed a higher impact on SAE, skewing towards a higher SHAP value. Ethnicity and race, specifically being of White race or Hispanic ethnicity, also showed substantial effects, followed by the medical history elements embedded within the eligibility criteria. Patients with a history of myocardial infarction or cerebrovascular accident within 12 months prior to randomization and interstitial lung disease were found to be significant in the model predictions, reflecting their potential impact on SAE.

**Figure 4.**
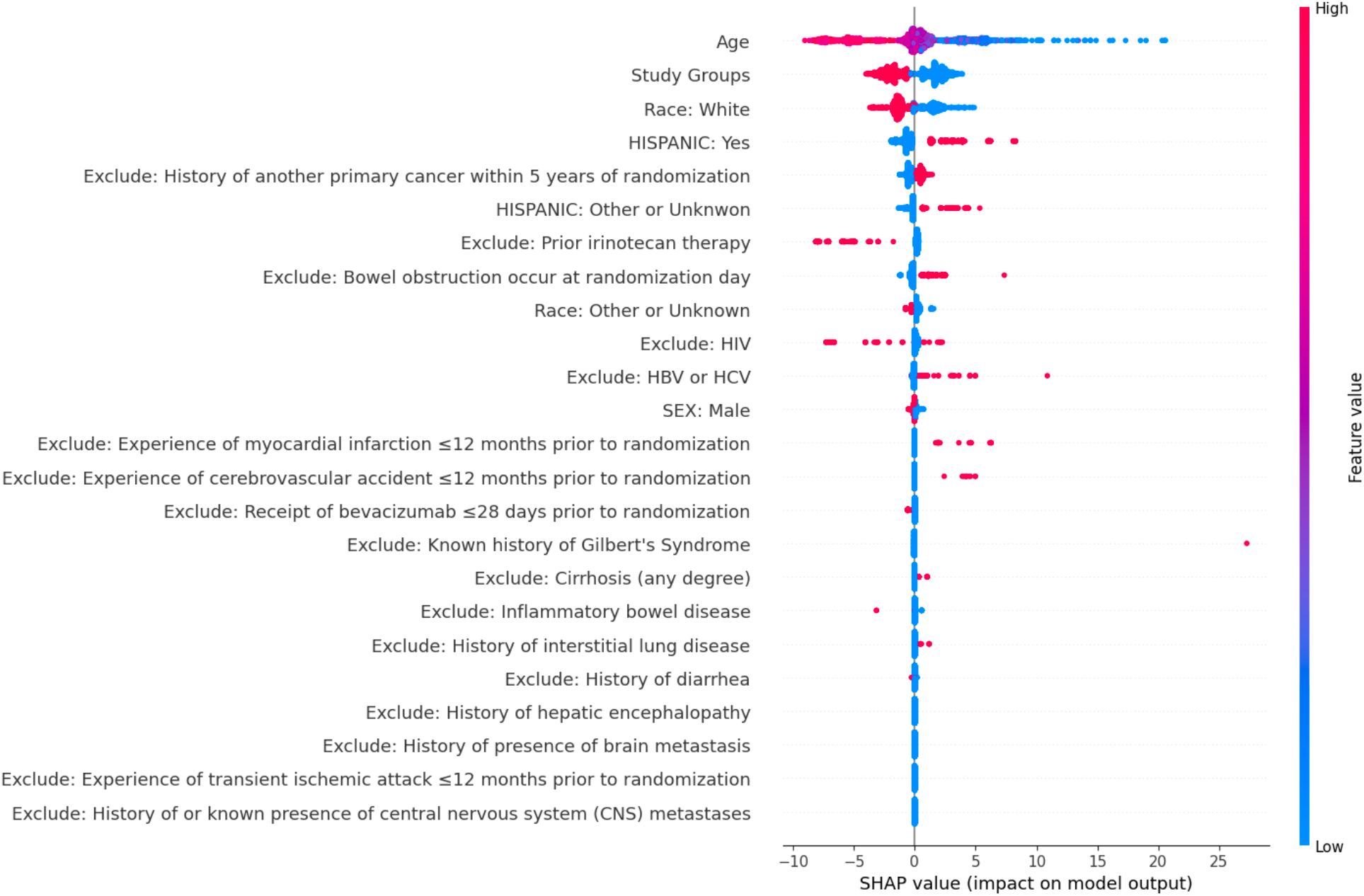
SHAP value for each patient.

## Discussion and Conclusions

This study’s findings contribute to a growing body of literature that seeks to understand how eligibility criteria can impact patient outcomes in clinical trials for CRC. Our investigation sheds light on the nuanced roles that demographic factors and eligibility criteria play in patient survival and the occurrence of SAEs.

The significant association between ethnicity and survival, particularly among Hispanic individuals, may reflect underlying social determinants of health or biological factors that warrant further study. Our results also challenge the notion that clinical trial outcomes are universally applicable, emphasizing the need for demographic diversity to ensure findings are reflective of the broader patient population. Furthermore, the elevated risk associated with a history of other primary cancers and recent myocardial infarctions points to a critical need for personalized medicine approaches in clinical trial design and patient care strategies. These medical histories are indicative of a patient subgroup with a more complex health profile, potentially requiring tailored interventions and monitoring. The observed increase in SAE risk with age and study group variances underline the need for age-appropriate dosing and treatment. This could have direct implications for clinical practice, suggesting that closer monitoring or adjusted treatment plans may be necessary for older CRC patients.

Interestingly, while some exclusion criteria, such as a history of interstitial lung disease and prior irinotecan therapy, showed no significant impact on the survival outcome, their influence on the risk of SAEs was noteworthy in the SHAP analysis. This discrepancy highlights the potential for eligibility criteria to have a differential impact on survival versus SAEs, suggesting that trial eligibility may need to be tailored to the specific outcome of interest. Our study has uncovered that specific medical histories, notably myocardial infarction and the presence of other primary cancers, significantly influence survival outcomes. This underscores a crucial finding: rigorous eligibility criteria may inadvertently sideline a critical portion of the broader CRC patient population, thereby constraining the external validity of our results. This raises ethical questions about the balance between patient safety and the inclusivity of trial populations. As we move towards an era where personalized treatments are becoming the norm, our study underlines the importance of re-evaluating current eligibility criteria to enhance the external validity of trial findings.

The limitations of our study include the retrospective nature of the analysis, reliance on EHR data, and the potential for unmeasured confounding factors. Despite these limitations, our results have important implications for future trial designs and highlight the need for a more granular understanding of how eligibility criteria affect trial outcomes.

In conclusion, the results of survival analysis, machine learning model, and SHAP value provide evidence that can inform the future design of CRC drug trials, with an emphasis on individualized patient selection that may enhance survival outcomes and reduce the incidence of SAEs. These insights will contribute to the optimization of clinical trial protocols and the advancement of patient care in colorectal cancer. Future research should focus on prospective studies to validate our findings and explore the mechanisms by which eligibility criteria influence patient outcomes. Additionally, incorporating patient-reported outcomes and quality-of-life measures into clinical trial designs could provide a more comprehensive understanding of the impact of eligibility criteria on patient experiences.

## Data Availability

All data produced in the present work are contained in the manuscript

## Acknowledgment

This study was partially supported by grants R01CA246418, R01CA246418-02S1, R21CA245858, R21CA245858-01A1S1, R21CA253394-01A1, R01AG080624, and R21AG068717 from the National Institutes of Health (NIH). The authors wish to thank the Cancer Informatics Shared Resource in the UF Health Cancer Center for data analytics support.

---

**Supplement Table 1.**
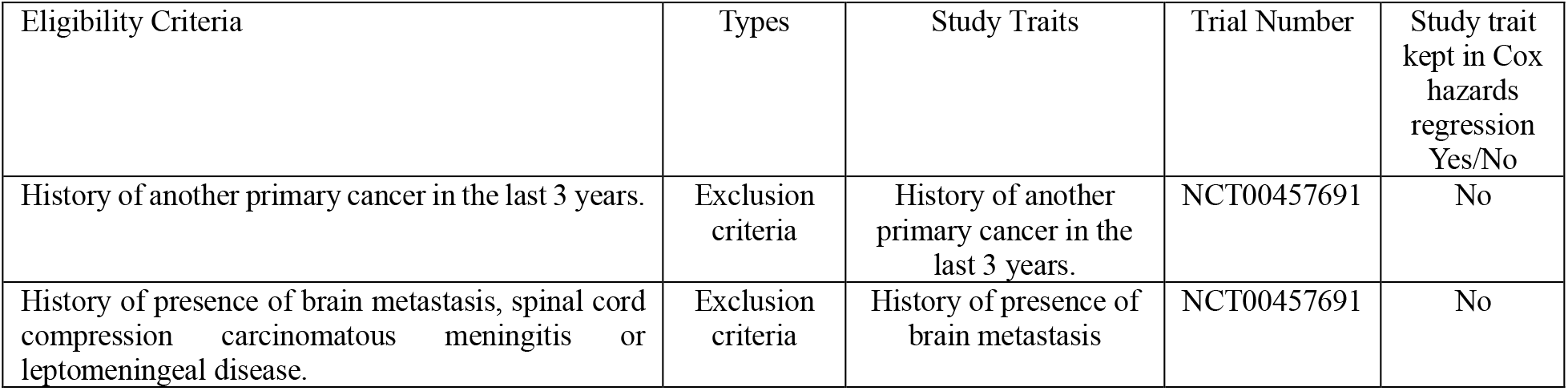
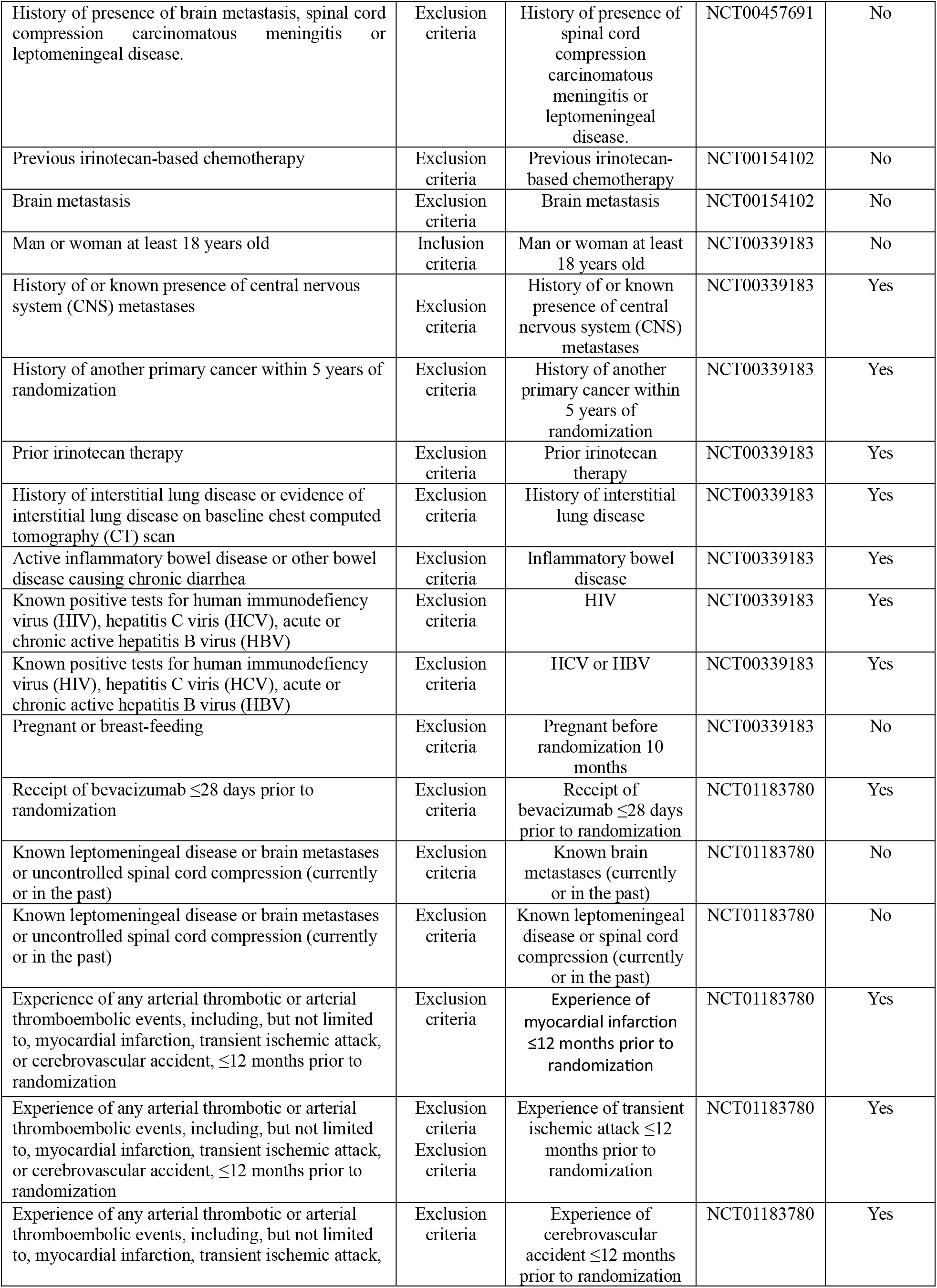
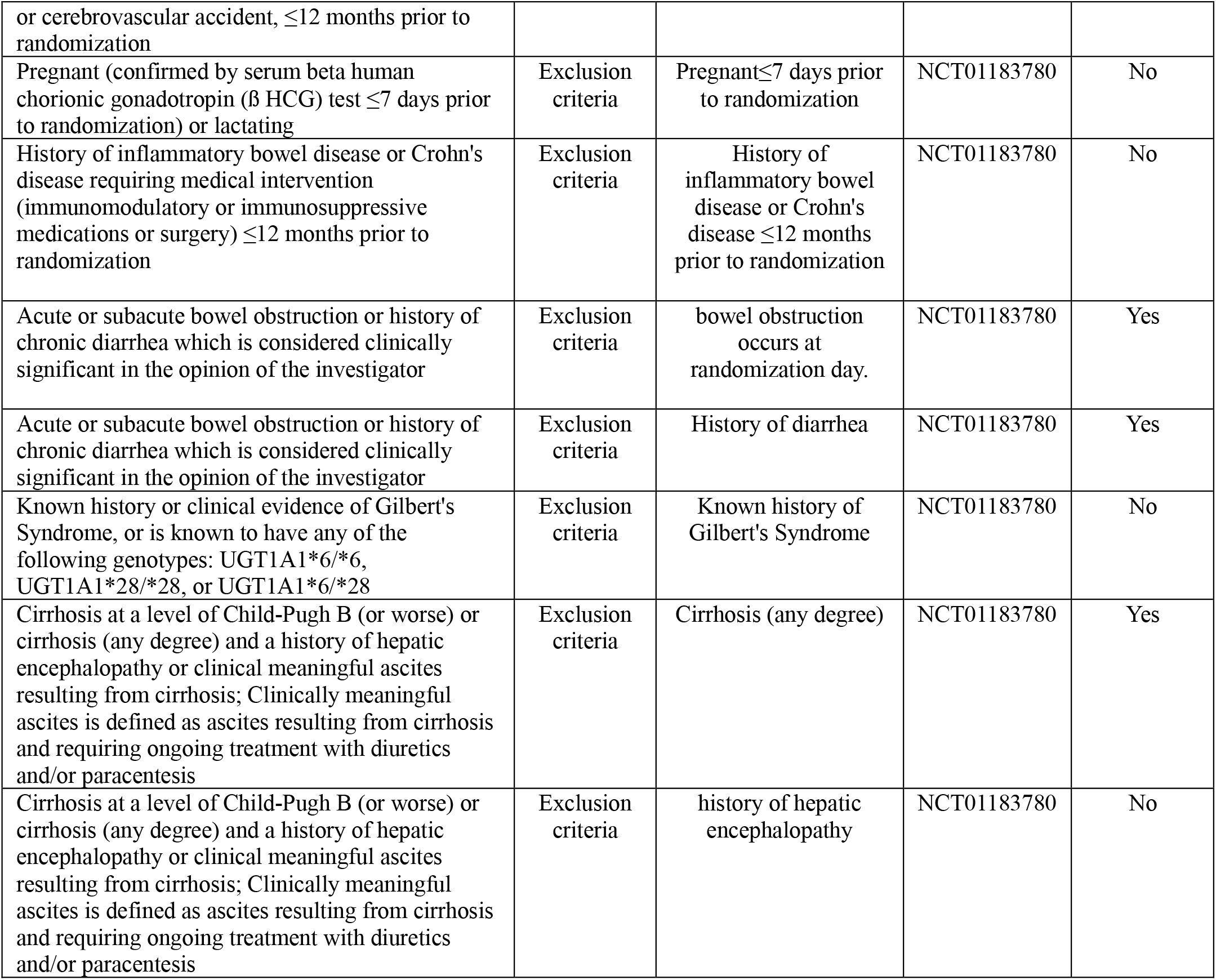
Computable eligibility criteria and decomposed study traits by trial.

